# Lies, Gosh Darn Lies, and Not Enough Good Statistics: Why Epidemic Model Parameter Estimation Fails

**DOI:** 10.1101/2020.04.20.20071928

**Authors:** Daniel E. Platt, Laxmi Parida, Pierre Zalloua

## Abstract

An opportunity exists in exploring epidemic modeling as a novel way to determine physiological and demic parameters for genetic association studies on a population/environmental (quasi) epidemiological study level. First, the spread of SARS-COV-2 has produced population specific lineages; second, epidemic spread model parameters are tied directly to these physiological and demic rates (e. g. incubation time, recovery time, transmission rate); and third, these parameters may serve as novel phenotypes to associate with region-specific genetic mutations as well as demic characteristics (e. g. age structure, cultural observance of personal space, crowdedness). Therefore, we sought to understand whether the parameters of epidemic models could be determined from the trajectory of infections, recovery, and hospitalizations prior to peak, and also to evaluate the quality and comparability of data between jurisdictions reporting their statistics necessary for the analysis of model parameters across populations. We found that, analytically, the pre-peak growth of an epidemic is limited by a subset of the model variates, and that the rate limiting variables are dominated by the expanding eigenmode of their equations. The variates quickly converge to the ratio of eigenvector components of the positive growth rate, which determines the doubling time. There are 9 parameters and 4 independent components in the eigenmode, leaving 5 undetermined parameters. Those parameters can be strikingly population dependent, and can have significant impact on estimates of hospital loads downstream. Without a sound framework, measurements of infection rates and other parameters are highly corrupted by uneven testing rates to uneven counting and reporting of relevant values. From the standpoint of phenotype parameters, this means that structured experiments must be performed to estimate these parameters in order to perform genetic association studies, or to construct viable models that accurately predict critical quantities such as hospitalization loads.

## Introduction

Infection^1,2^, transcription and replication^3,4^ by SARS-COV-2 involves a number of rate limiting interactions with host cells that are likely to be modulated by mutations in cellular as well as viral genes. At the same time, phylogenetic analysis shows geographic specificity^5,6^, indicating that geographic regions may show specific exposure to distinctive SNP combinations, or viral haplotypes, in SARS-COV-2. This specificity suggests a benefit to exploring relationships between duration of the prodromic phase, proportions of asymptomatic cases^7^, proportions of severe cases, rates of recovery, among other infection attributes^8^, that define temporal progression of compartmental epidemic models, starting with SIR (Susceptible-Infected-Recovered) models^9^. Beside host and viral genetic impacts, other aspects driving SARS-COV-2 rates are population specific and demic, such as the impact of age on both asymptomatic and mild cases, as well as the proportion of severe and critical cases. Other aspects include normal social distance, and how effectively social-distancing rules have been followed. Hospital survival may also reflect impacts of some genetic susceptibility, presence of comorbidities (Hypertension, Diabetes, Asthma, lung disease, obesity and others yet to be identified) as well as the level of stress on the region’s medical facilities and medical staff.

In this paper, we seek to identify the limitations of using compartmental models to estimate or test hypotheses concerning parameters governing the growth of SARS-COV-2 epidemics. We also seek to investigate what type of epidemic variable tracking is necessary to effectively quantify the parameters that are suitable for hypothesis testing at the level of environmental exposure in epidemiological studies.

## Methods

Compartmental models count individuals at different stages of progression of a disease, where each stage of progression is marked by an event that has a well-defined rate. For example, from time of infection to the time the person can transmit disease has a time distribution, that, for enough people in the compartment, will tend to center on an average by the central limit theorem for large enough samples drawn from any given distribution. There is evidence that COVID-19 presents symptomatic cases and asymptomatic cases, with asymptomatic cases^10–12^ less likely to be identified and isolated^13–17^. There is an incubation period after infection that lasts until the incubating individuals become infectious. There has been some early estimates based on confirmed cases^13,18^ with more evidence of pre-symptomatic transmission being noted^19,20^ yielding faster incubation. Incubation partly accounts for the observed lag when social distancing or other viral spread prevention policies are imposed. For the most part, infections appear to be transmittable prior to overt symptoms, allowing for a pre-symptomatic period that may convert to symptomatic. At the same time, some of those asymptomatic people remain asymptomatic until they are non-contagious^11^. Patients may still be infectious for several days after symptomatic recovery. Symptomatic patients likely to be hospitalized are hospitalized more quickly than non-hospitalized patients recover. Hospitalized patients in ICU or required immediate ventilation tend to experience a longer time to recovery than non-hospitalized patients. Those that stay on the ventilator for long periods tend to have a high mortality rate, and may stay on the ventilator for many weeks prior to dying.^8^

A compartmental model that captures the conditions status and durations count susceptible population members *S*, incubating *E*, infectious asymptomatic *I*_*A*_, infectious symptomatic *I*_*S*_, infectious people who will be hospitalized *I*_*H*_, those hospitalized who recover *I*_*HR*_, and hospitalized leading to mortality *I*_*HM*_. Recoveries are *R*, and mortalities are *R*_*M*_. The time from exposed to infectious is *α*^−1^, where *α* is partitioned into contributions to asymptomatic infectious *I*_*A*_, symptomatic infectious *I*_*S*_, and infectious that will be hospitalized *I*_*H*_, so that *α* = *α*_*IS*_ + *α*_*IA*_ + *α*_*H*_. Total removal time among asymptotic infectious is 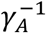, with a fraction *ζ* going to infectious symptomatic. Infectious symptomatic removal time is 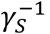. The period prior to hospitalization is (*α*_*HR*_ + *α*_*HM*_)^−1^. The rate that the proportion that recovers is *α*_*HR*_, and that which dies is *α*_*HM*_. The model equations, reflecting an underlying Markov chain, expressing these connections and rates are:

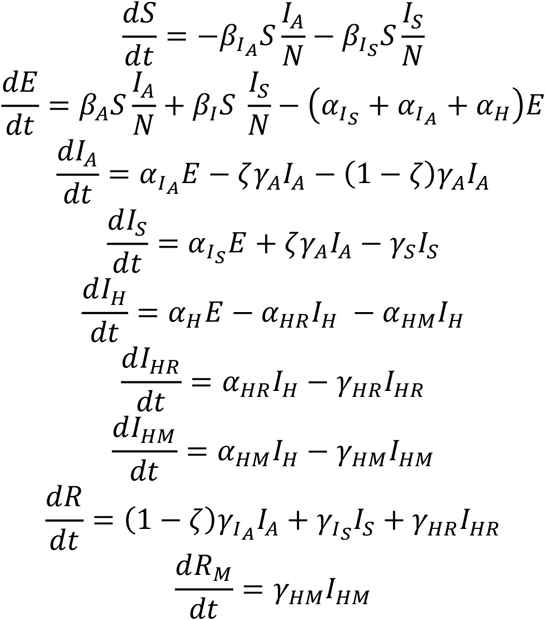

where

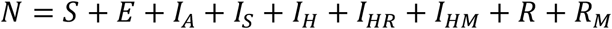

Note that 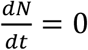, indicating conversions of all individuals in the system are accounted for. Parameter values derived from publications are listed in Table 1.

**Table 1.** Published times for compartmental conversions, proportions, and derived rates.

| Parameter | Times | Value | Notes |
| --- | --- | --- | --- |
| $\alpha$ | 5.1 (4.5-5.8) days<sup>22</sup>, 5.2 (4.1 – 7.0)<sup>19</sup> 5.2 (3.78-6.78)<sup>18</sup> – 3.95(3.01-4.91)<sup>18</sup> | 0.196, 0.25 | $\alpha = \alpha_{IS} + \alpha_{IA} + \alpha_H$ |
| $\eta_S$ | 30.8%<sup>23</sup>, 20.6% (23-33%) – 40%(36-44%)<sup>12</sup> | 0.3 | $\alpha_S = \eta_S \alpha$ |
| $\eta_A$ | 49.2% (by total), 68.7 | 0.492 | $\alpha_A = \eta_A \alpha$ |
| $\eta_H$ | 20%<sup>8</sup>, 1.3% | 0.2, 0.013 | $\alpha_H = \eta_H \alpha$ |
| $\eta_{HR}$ | 69%<sup>8</sup> | 0.69 | $\alpha_{HR} = \eta_{HR}(\alpha_{HR} + \alpha_{HM})$ |
| $\eta_{HM}$ | 31%<sup>8</sup> | 0.31 | $\alpha_{HM} = \eta_{HM}(\alpha_{HR} + \alpha_{HM})$ |
| $\alpha_S$ | | 0.0588, 0.075, 0.172 (est) | $\alpha_S = \eta_S \alpha$ |
| $\alpha_A$ | | 0.0964, 0.123 | $\alpha_A = \eta_A \alpha$ |
| $\alpha_H$ | | 0.0392, 0.05, 0.00325 (est) | $\alpha_H = \eta_H \alpha$ |
| $\alpha_{HR} + \alpha_{HM}$ | 7 days<sup>8</sup> | 0.143 | |
| $\alpha_{HR}$ | | 0.09867 | |
| $\alpha_{HM}$ | | 0.04433 | |
| $\beta_A$ | | | |
| $\beta_I$ | | | |
| $\zeta$ | 20.8 %<sup>11</sup> | 0.208 | |
| $\gamma_A$ | 14 days<sup>8</sup>, 9.5days<sup>11</sup> | 0.0714, 0.105 | |
| $\gamma_I$ | 14 days<sup>8</sup>, 9.5days<sup>11</sup> | 0.0714, 0.105 | |
| $\gamma_{HR}$ | 31 days<sup>8</sup> | 0.0323 | |
| $\gamma_{HM}$ | 42 days<sup>8</sup> | 0.0238 | |

The rate of infection for a susceptible individual depends on the probability that an infectious viral load is transferred, multiplied by the rate of encounters a susceptible individual has. The encounters can involve: other susceptible individuals, or symptomatic infectious people, which as a group tends to be isolated with a corresponding depressed rate of encounters 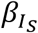, and undetected asymptomatic infectious people whose interaction rate 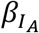 is substantially higher, subject to social distancing regulations. The fraction of infectious symptomatic individuals that a given susceptible individual may encounter is 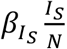, and the total number of susceptible individuals exposed to infectious symptomatic cases is 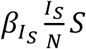. Likewise, that for asymptomatic cases, the rate of symptomatic infections is 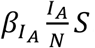. These terms drive the creation of new infections in the population. The force of the symptomatic group is the coefficient of *I*_*S*_, or 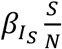. The number of the susceptible group that an individual can infect over their entire period of infectiousness is the reproduction number 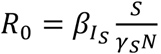, and similarly for the asymptomatic infectious group. These numbers primarily drive the rate of growth of the infection in the population, which early in the expansion is measured by the doubling time.

Early in the evolution of the infection, which may be defined as when *N* − *S* ≪ *N*, the variables immediately involved in the feedback loop determine the rate limiting step. Therefore, identifying

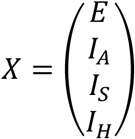

and

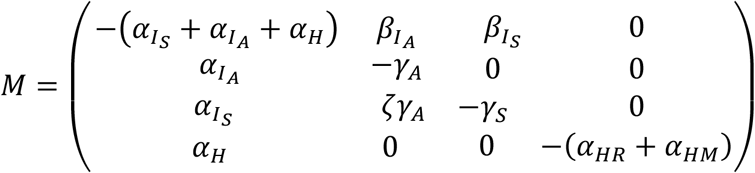

the equation governing the system in this regime is

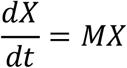

This has solutions of the form *X*(*t*) = *e*^*Mt*^*X*(0). The *M* may be diagonalized by a matrix *U* so that *U*^−1^*MU* = *K*, for 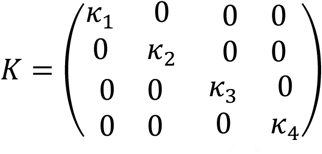. Then 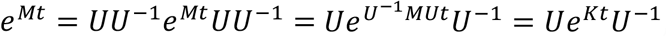, and 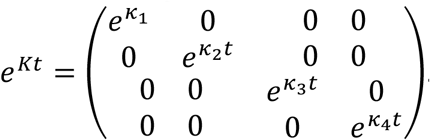. Since *MU* = *UK*, Each of the columns of *U* are eigenvectors *u*_*j*_, where *Mu*_*j*_ = *k*_*j*_*u*_*j*_. This is an eigen equation, where the *k*_*J*_s determine the time rate of exponential growth or decay with doubling time 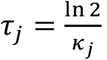, and the eigenvectors represent the linear combinations of *E, I*_*A*_,*I*_*S*_, and *I*_*H*_ that grow or decay with the that eigenvalue. The combinations of eigenmodes is determined by initial conditions. The leading eigenvalue will dominate with exponential growth yielding fixed proportions of each of the *E, I*_*A*_,*I*_*S*_, and *I*_*H*_ to each other. The other terms turn out to identify rates related to the delay time for the system to respond to changes in distancing policy due to incubation time, to imbalances between symptomatic and asymptomatic patients, and to the decay of *I*_*H*_.

Data from New York State were obtained from The COVID Tracking Project^21^.

## Results

Testing in New York State, starting on 03/04/2020, labeled as day 1. On 3/13, day 10, NY State received permission to contract for its own SARS-COV-2 testing. Statewide “distancing” started on 3/20, day 17, with the signing of the “New York State on Pause” bill. Prior to that, local jurisdictions had already been imposing local ordinances against assembly, and started closing schools.

Figure 1 shows the cumulative total testing and positive test numbers indexed by day. Testing has been driven by tracking contacts of discovered cases which is reflected heavily in the close alignment of total tests and positive tests. On 3/13, the total number of tests increased from 308 to 3200, with surges to the 5,000 level, then 7,000, then 14,000 showing rapid subsequent growth.

**Figure 1.**
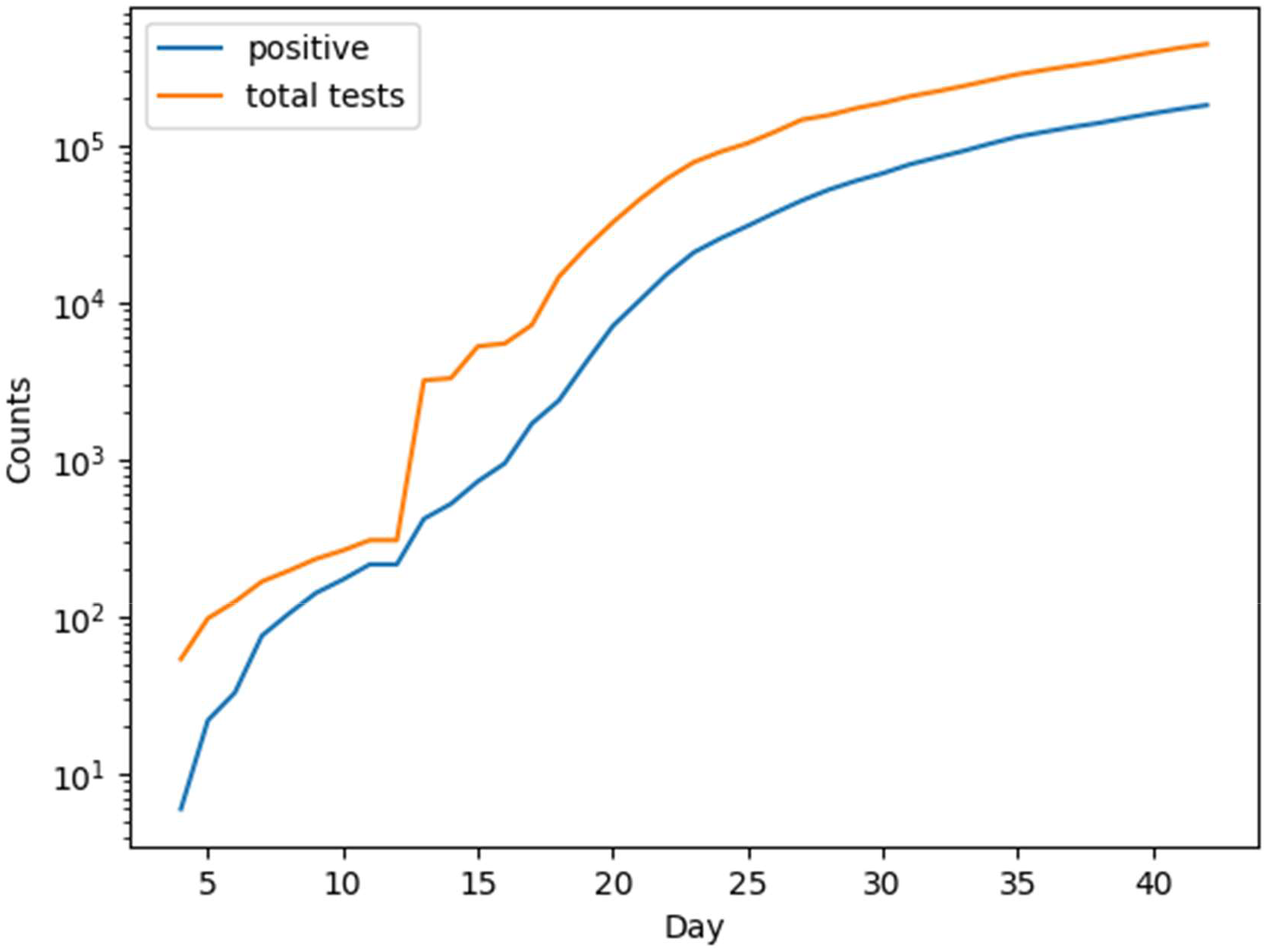
Levels of total testing and positive cases identified in New York State.

Early in the testing, from day 1 to 19, the rate of growth of positive cases was *κ* = 0.3519 ± 0.01390, corresponding to a doubling time of 1.97 ± 0.08. From day 20 to day 30, the rate of growth of positive cases was *κ* = 0.2027 ± 0.0076, corresponding to a doubling time of 3.42 ± 0.13. These numbers suggested very high rates of contagious transmission. These doubling times were reported by Governor Cuomo in some of his earliest briefings.

If, as tracking numbers increased, testing surveillance was broad enough to pick up community spread individuals proportional to total numbers of tests applied, then the proportion of positives from the tests may reflect population rates. However, if rates are tightly limited to immediate known cases, then the reported positives will be a better estimate of underlying population, since the fraction of those seeking medical assistance should be proportional to the exposed number in the population. When available tests increased, the apparent rate grew substantially. Therefore, infected population growth may be more closely reflected in the fraction of positive results normalized by total number of tests applied, in spite of very highly biased sampling selection. For a given proportion of ill patients who seek help, this should track with the fraction of the population who is ill. However, this may be subject to growing awareness of the population to get help with COVID-19 infections.

First, consider the idea that tests may be broad enough to sample spread. When test numbers were low, the likelihood that targeted testing would reflect the general population was also low and sampling uncertainties large. Therefore, a lower bound on testing levels was applied. This cut samples prior to 3/20. Later, test ratios started to demonstrate a downwards bend. This shoulder was cut for samples beyond 3/30. New York doubling time was estimated from a *χ*^2^ regression between the log of positive test ratios versus time, yielding *κ* = 0.0471 ± 0.0095 with a doubling time of 14.7 ± 3.0 adjusting for testing counts. In the alternative scenario, positive samples reflect the proportion of symptomatic patients seeking medical aid, a possibility since the testing was so closely tied to diagnosed patients plus contact surveillance. A regression was performed on the cumulative positive counts shown in Figure 2c) yielding *κ* = 0.1170 ± 0.0021 per day, with a doubling time of 5.9 ± 0.1 days.

**Figure 2.**
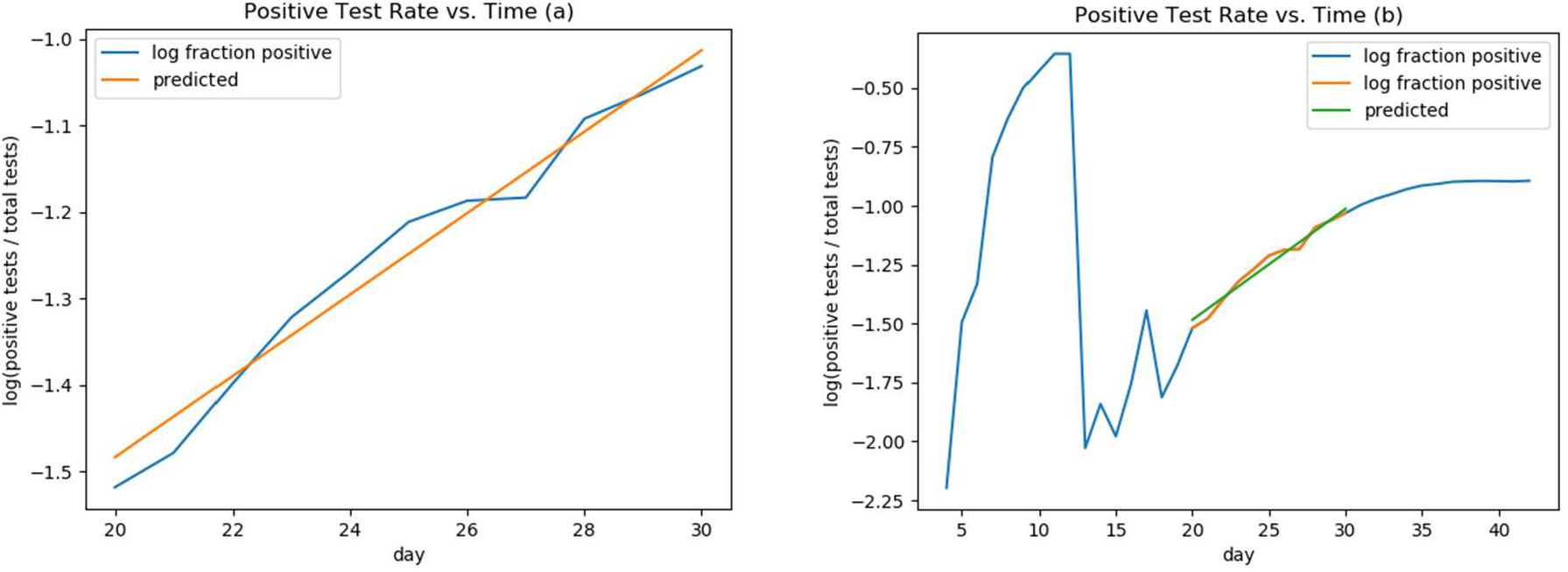
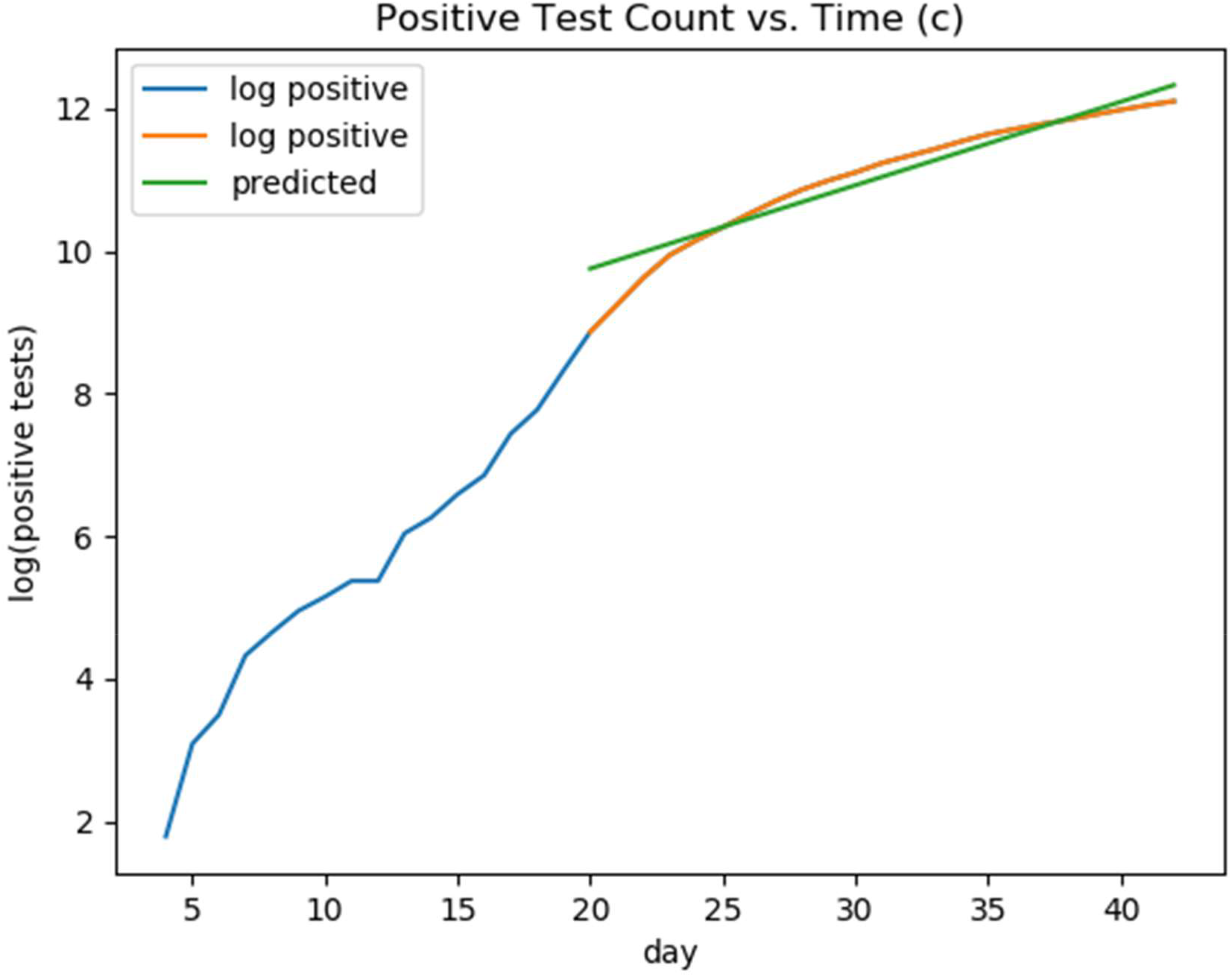
Log-linear *χ*^2^ regression estimate of *κ* from New York State growth of fraction of positive tests. a) linear regression representing a segment of the positive test rate vs. time; b) linear regression from a) represented within the entire test rate vs. time dataset; c) is a fit to the log of the positive test count vs days starting at 20 days.

Taking guidance from Table 1, values *α* = 0.25, 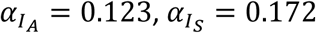, *α*_*H*_ = 0.00325, *α*_*HA*_ = 0.09867, *α*_*HM*_ = 0.04433, *ζ* = 0.3, *γ*_*A*_ = 0.0714, *γ*_*S*_ = 0.0714, *β*_*A*_ = 0.4748, and *β*_*I*_ = 0.1071 yields a doubling time close to New York State from Figure 2c. Figure 1 presents a log-linear plot of the growth of the complete model equations integrated numerically using solve_ivp() employing RK45 from scipy, clearly showing that the early growth is dominated by a leading exponential mode. The early lead-in shows the effects of decaying modes as the initial conditions converge to the fixed ratios of the leading eigenmode components. The leading eigenvalue is *κ* = 0.1171, yielding a doubling time of 5.9 days, with eigenvector *u* = (0.6457 0.4212 0.6369 0.0081). The component associated with incubation decay is *κ* = −0.483, associated with a response to policy change delay half-life of 1.4 days. Its eigenvalue is *u* = (0.8940 −0.2671 −0.3596 −0.0085). The eigenvalue *κ* = −0.0750 with half-life of 9.2 days is associated with deviations between *I*_*S*_ and *I*_*A*_ from the dominating growth eigenvector, and has an eigenvector of *u* = (−0.0064 0.2172 −0.9761 −0.0003). The last eigenvalue is *κ* = −0.143, associated with the decay of *I*_*H*_ from equilibrium values with eigenvector *u* = (0 0 0 1).

Figure 3 shows a log-linear plot of the rate-limiting variables for a numerical integration of the entire system of differential equations. The pre-peak segment shows a clear view of how the system is dominated by the leading exponential eigenmode of the growth, including the proportions between variables represented in the eigenvector of the leading eigenvalue, which determines the slope.

**Figure 3.**
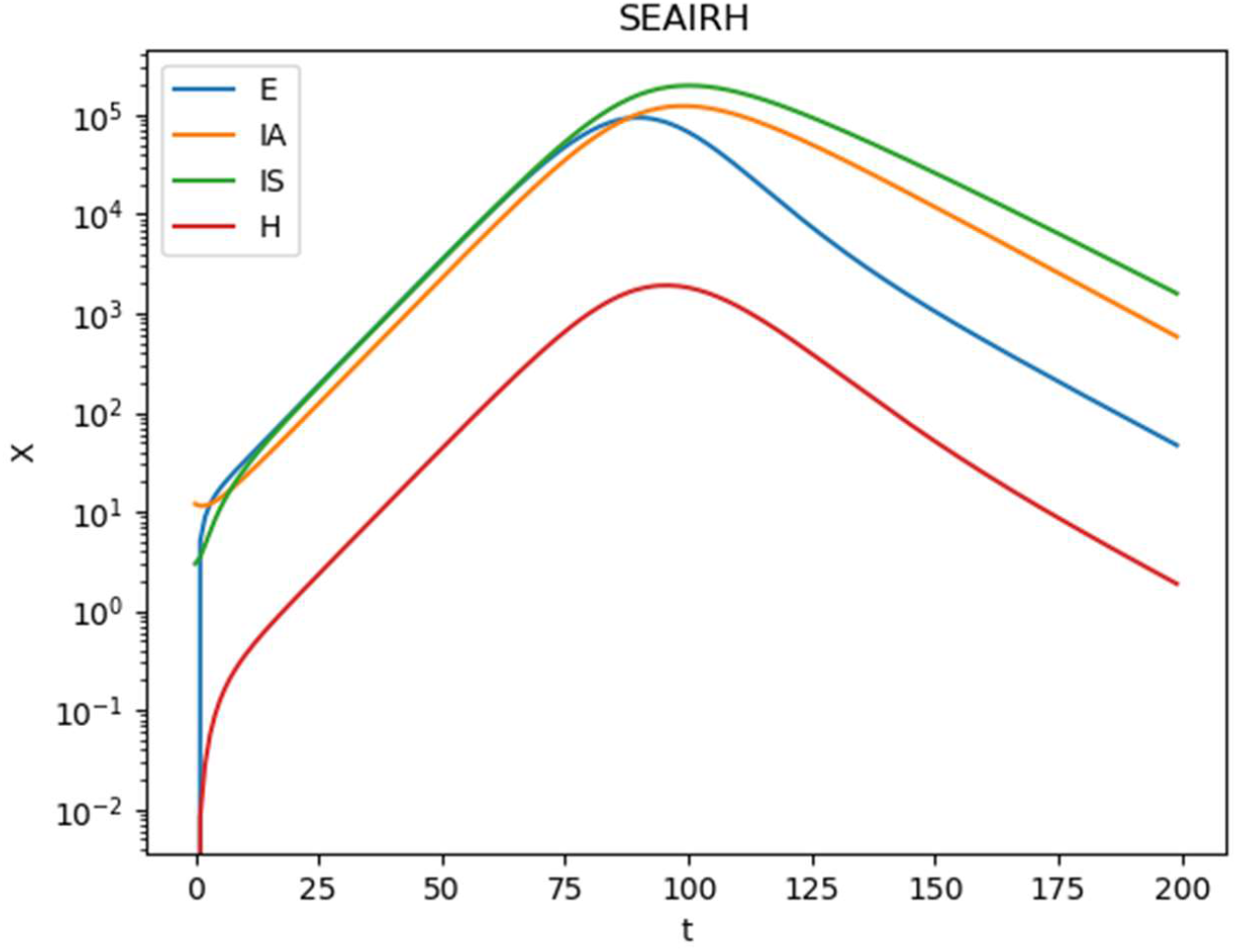
Log-linear plot of rate-limiting variables in the full system of equations integrated numerically.

Figure 4 shows the evolution of the system variables in a linear-linear plot. The lags in the peak variables shown in figure 4a identify the peak pulse through the system of linear equations. The “est” entries in Table 1 for *α*_*H*_ represent values commensurate with (but not a fit to) the NY hospitalization levels^21^. They are a factor of 12 smaller than those fitting the Wuhan hospitalization rate^8^. As such, it is clear that the impact of COVID on features such as progression to hospitalization, response to treatment for symptomatic patients, whether patients are identified in time to stop progression to serious or critical stages may impact survivability. The model predicts 3294 fatalities per million, peak recovering hospitalizations of 3347 on day 111, and peak mortality hospitalization (primarily long-term ventilator load) of 1732 on day 114. Figure 4b includes susceptible *S* and recovered *R* variables. The range of variation of these variables appears to dwarf the fraction of the population that is incubating, infected, or involved with hospital load. One feature of the equations is that the rate of flow of individuals through a compartment may not be reflected in the total number in the compartments at any given time, even at their peaks. At the end, these rates would leave 24,738 per million uninfected and susceptible, with 971,967 recovered per million.

**Figure 4.**
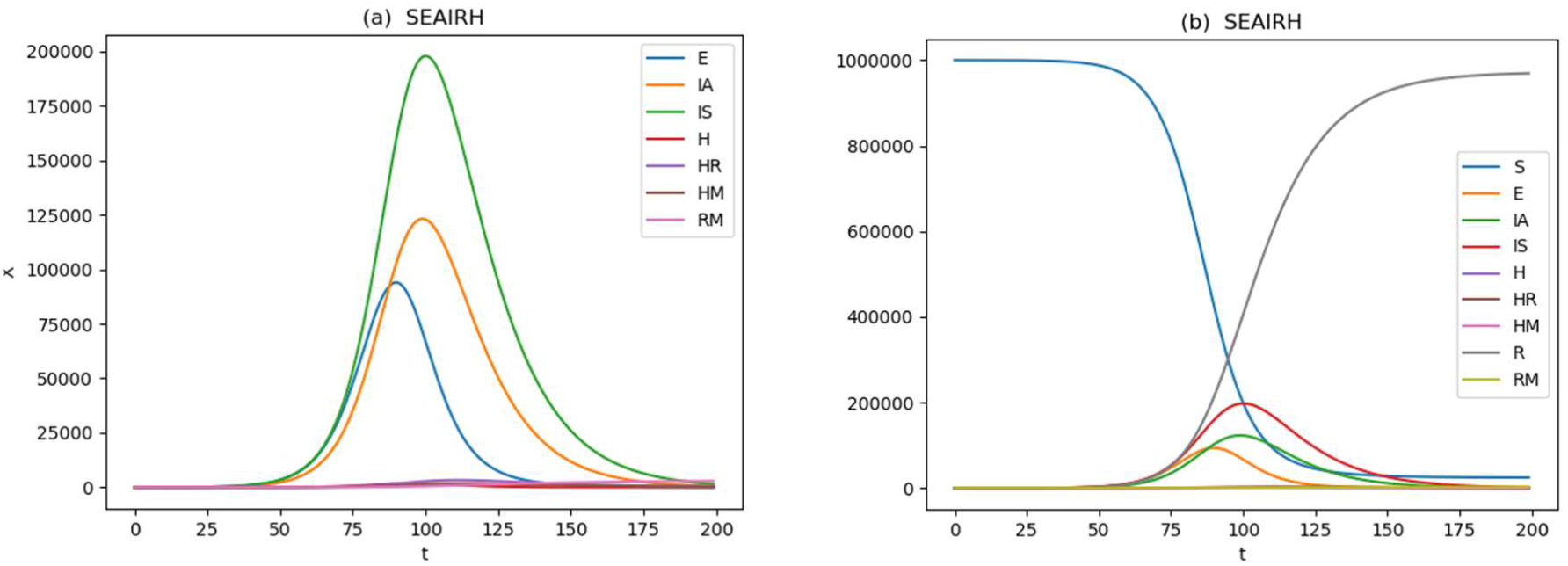
The evolution of the model given the apparent doubling time represented by the regression in Figure 2. The peaks in variables in (a) show lagging as the compartments move through their sequence. The susceptible and recovered variables are included in (b).

The difficulty in understanding how the testing protocol impacts estimations of rates is illustrated in the New York State rates shown in Table 2. Considering cases as a representative sample of a fixed proportion of the infected population argues for computing a rate based on cumulative cases. If, on the other hand, the testing generated a random sample of the broader population, more testing would identify more individuals simply because there were more tests. If so, the proportion of positives to total tests may be a closer approximation to the population, and the total positives would be proportional to the square of the actual proportion of diseases, resulting in a doubling of *κ*. That seems to be roughly what was observed between the two New York State regressions. On the other hand, cumulative rates for two other jurisdictions, Lebanon and New South Wales, Australia, show rates similar to each of the two New York State numbers. And while the New York State proportional model gives an expected factor of 2 in the rate, it is the cumulative rate that more closely resembles the growth and peak in New York, not the relative proportion rate. More, the shifts in test availability and distancing initiation are all visible in the New York data, which contributes to the difficulty even of identifying exponential growth regimes, much less identifying an exponential rate that constrains the available model parameter space.

**Table 2.** Exponential growth rates, corresponding doubling times for various populations and measurements given available data.

| Region | $\kappa$ | $T_d$ (days) |
| --- | --- | --- |
| **New York State<sup>21</sup>**<br>**(cumulative cases)** | $0.1170 \pm 0.0021$ | $5.9 \pm 0.1$ |
| **New York State<sup>21</sup>**<br>**(relative frequency)** | $0.0471 \pm 0.0095$ | $14.7 \pm 3.0$ |
| **Lebanon<sup>24</sup>**<br>**(cumulative cases)** | $0.05998 \pm 0.00786$ | $11.6 \pm 1.5$ |
| **Australia New**<br>**South Wales<sup>25</sup>**<br>**(cumulative cases)** | $0.1984 \pm 0.0153$ | $3.5 \pm 0.3$ |

## Discussion

One of the major goals of epidemic modeling is to predict mortality and resource load on community medical facilities: how many beds, how many ventilators, how much pharmaceuticals, among other resources will be needed to get through the epidemic. Early epidemic growth for this system is dominated by the largest eigenvalue of 9 coefficients governing the rate-limiting variables. This eigenvalue determines the doubling time of the growth, and imposes one constraint on those coefficients; the eigenvectors impose three more constraints on the system, leaving five coefficients undetermined. Essentially, all of the rate-limiting relevant epidemic variables grow at the same rate maintaining fixed ratios. However, as they near peak, the variable trajectories become more differentiated, with lagging or leading peaks emerging as the impact of 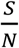 filters through the system of equations. However, at peak, it is already too late to allow time to acquire and deploy needed resources to hospitals and clinics. By itself, the trajectory of these models in pre-peak growth offer little hint as to final needs. Further, there are a number of combinations of parameters that would yield the same leading eigenvector and eigenvalue.

More so, the parameters that govern these epidemic models tend to reflect physiological rates of how the disease expresses itself in individuals, as well as effects that are moderated by demic characteristics. Examples are age structure in the population, which impacts both asymptomatic cases^11,26^ and severity of disease^8^. Identification of asymptomatic cases has been problematic since testing protocols tended to require symptoms, or contacts with known infected people. One case in California went untested for 10 days because she had no known contacts. Cases that advance to severe or critical depend on other factors, such as treatment modalities prior to development of advanced symptoms. The rate of transmission depends on physiological parameters as well as normal social distance and social distancing response to an epidemic, how public institutions such as schools are run, how grocery shopping interactions are handled, whether known infections are isolated and other factors specific to each community. Given how widely these parameters may vary from population to population, how they vary: how they depend on the geographically specific dominating SARS-COV-2 lineages dominant within a given geography^5,6^, and how they depend on behavioral, social, age structure, and other factors of a population, it is worth seeking whether and how these factors relate to the expressed epidemic model rate parameters as phenotypes.

Since the problem of identifying rate limiting parameters prior to peak is underdetermined, these rates must be determined elsewhere. Most statistical reporting does not provide nearly enough information to extract these factors, even at an environmental (quasi-) epidemiological experimental design standards. Further, jurisdictions are applying tests to try to identify new cases that are related to other identified cases through contact. The “enrollment protocol” was not designed to understand the spread in the population, but rather to try to identify patients and remove them from circulation by isolating them. More and broader testing is applied as test kits become more available. Test kits may not be uniform with loss of sensitivity depending on the stage of the infection and/or the type of swab taken (Nasal, nasopharyngeal or sputum). From jurisdiction to jurisdiction, testing and reporting protocols vary, making it difficult to compare jurisdictions, or even the same jurisdiction to itself from day to day. The rate of growth and doubling time may reflect availability and levels of testing more than the actual disease in the population.

Perhaps the best way to acquire the necessary parameters would be a prospective longitudinal study cohorts in multiple jurisdictions. Enrollment should be randomized, reflect regional characteristics such as sex and age, and the criteria should be shared across populations participating in the study. During the course of the study, subjects will be monitored for changes in status a) from susceptible to incubating recording dates of exposure (if possible), b) to infectious (symptomatic or asymptomatic, with a clearly defined standard for determining possible “infectious” condition) conversion and dates, c1) for asymptomatic to symptomatic conversions and dates or c2) recovery dates, d) symptomatic to recovery conversion dates, or e1) hospitalization dates, e2) recovery from hospitalization dates, e3) ICU admission dates, e4) ICU recovery date, e5) ventilator treatment start date, e6) ventilator recovery date, e7) date of death. A record of how each subject moves through the model compartments, together with time distributions, can provide phenotypic parameters that modelling alone cannot, offering insight into the biology, response of the disease to medications, comorbid conditions, demic characterizations, and other features relevant to the impact of COVID-19.

Further, these parameters provide a uniform basis for comparisons between populations necessary for complete model constructions that yield distributions of trajectories and confidence intervals for timing and peak loads, and which can provide a full epidemiological exploration of how individual subject phenotypes respond to environmental, genetic, comorbid, and behavioral factors that may yield valuable information for biological, clinical and pharmaceutical development. As such, these models may be used to test and verify measurements of physiological parameters, and to identify evidence whether some factor strong enough to generate deviations is missing.

## Conclusions

A response to an article in Nature^27^ stated: “A well-known lawyer, now a judge, once grouped witnesses into three classes: simple liars, damned liars, and experts. He did not mean that the expert uttered things which he knew to be untrue, but that by the emphasis which he laid on certain statements, and by what has been defined as a highly cultivated faculty of evasion, the effect was actually worse than if he had.” The statement was applied to the specific issue of expert forensic testimony. The statement has been restated as “lies, damn lies, and statistics.” The message serves as a warning that statistics collected for certain purposes may not be suited to other purposes. That unsuitability does not reflect any attempt at obfuscation. Specifically, in this case, the use of testing, positive test counts, etc are tilted towards identifying patients who are likely to have specific treatment needs, and to try to identify contacts to stop epidemic spread. These uses render the reported statistics problematic for modeling. Physiological parameters based on identified patients may be biased in terms of the patients who were identified, and the methods by which they were identified. Further, protocols shifted as previously unrecognized community spread and asymptomatic individuals were recognized to be significant contributors to viral spread.

Finally, modeling not only can provide important information planners need for capacity loads, but models can also test whether the parameters as understood describe how the disease behaves in a population. A failure to predict may indicate an important factor in the disease’s behavior that had not been recognized. In order for this to work, a more formally structured prospective cohort study, with adequate annotations of pharmaceuticals, comorbidities, and other factors, is likely the best way to ensure all the rates are measured on a consistent footing throughout the course of the epidemic.

## Data Availability

Data are freely available at cited websites

https://covidtracking.com/

https://www.wolframcloud.com/obj/resourcesystem/published/DataRepository/resource

https://www.moph.gov.lb/maps/covid19.php

